# Health-Related Quality of Life Among Patients Suffering from Myopia or its Long-Term Consequences: A Cross-sectional Survey Protocol

**DOI:** 10.1101/2025.10.21.25338448

**Authors:** Sarah Dewilde, Julie Boelen, Kun Shi-van Wielink

**Affiliations:** Services in Health Economics (SHE), Brussels, Belgium; Santen Pharmaceuticals, Amsterdam, The Netherlands

## Abstract

**Background:** Myopia, commonly referred to as near-sightedness, is the most common ocular disorder worldwide. If left untreated, high myopia in children can lead to serious eye diseases in adulthood. To mitigate the risk of long-term complications, it is essential to implement interventions aimed at slowing the progression of myopia. To evaluate the value of such interventions, cost-effectiveness models need to be developed. In cost-effectiveness models, health-related quality of life (HRQoL) is often expressed as a “utility value”. However, the literature on utility value in relation to myopia severity and its complications is very limited.

**Objective:** This study aims to estimate HRQoL and utility values of myopia in both childhood and adulthood, to provide input for cost-effectiveness models and ultimately enhance access to myopia treatments.

**Methods:** This is a cross-sectional study that will invite patients from hospitals to complete an online survey across seven countries (United Kingdom, France, Germany, Italy, Spain, Denmark, and Sweden), with a minimum of 50 patients from each country (n=350). Participants include proxies of children diagnosed with myopia and adults suffering from myopia and related long-term complications. The survey will include closed-ended questions for validated instruments that assess HRQoL, functioning, and healthcare utilization. These include the EQ-5D-Y-3L, EQ-5D-5L, with bolt-on for “vision”, PedEyeQ, and NEI-VFQ-25.

**Results and dissemination:** Data collection is scheduled to start in June 2025, with results expected by Q4 2025. Results aimed to be published as open access in peer-reviewed high-impact journals.

**Conclusion:** The data collected will be used to evaluate the impact of myopia on HRQoL, inform health economic models, and enhance access to myopia treatments.

## 1. Background

Myopia, commonly known as near-sightedness, is a refractive error of the eye that causes distant objects to appear out of focus. It is the most common disorder worldwide, with an estimated prevalence of 3.4 billion people by 2030^1,2^. If left untreated, high myopia in children can lead to serious eye diseases in adulthood, such as retinal detachment, cataract, myopic macular degeneration, open-angle glaucoma, and, in severe cases, visual impairment or blindness^1,3-5^.

To reduce the risk of these long-term complications, interventions that slow the progression of myopia, rather than simple optical correction, are required ^6^. To assess whether the value of such interventions justifies their cost and whether they should be incorporated into routine care or reimbursed, budget impact analysis and/or cost-effectiveness analysis must be performed. Cost-effectiveness analysis combines interventions effectiveness with its associated healthcare costs (i.e., treatment costs, visits to general practitioners and specialists, medical interventions, and hospitalizations) but also societal costs such as productivity loss. HRQoL expressed as quality-adjusted life years (QALYs), is often the standard measure of effectiveness used in cost-effectiveness analyses.

Myopia can impact various aspects of a child’s daily activities, including school performance, sports, and social interactions. Understanding these effects is crucial for evaluating the overall effectiveness of interventions aimed at improving the HRQoL for children with myopia. Additionally, the long-term complications of myopia can significantly impact adults’ daily functioning, mental well-being, and independence.

Patient satisfaction with current treatments plays another key role in evaluating treatment effectiveness. Identifying challenges or side effects, such as discomfort and dry eyes associated with existing treatments, will help improve myopia management options for children. Given the differences in health care systems and reimbursement structures across Europe, country-specific data on health-care resource utilization is essential for an accurate assessment of the cost-effectiveness of myopia interventions.

In cost-effectiveness models, HRQoL is often expressed as a “utility value”. This numerical representation reflects a patient’s preference for a particular health state with values ranging from 0 (representing death) to 1 (representing perfect health). The values help quantify the impact of different health conditions or treatments on overall well-being. However, the literature on utility values by myopia severity and for its complications is very limited^7,8^.

This survey aims to estimate utility values for myopia and its related complications in both childhood and adulthood. By assessing the HRQoL in these individuals, we can gain insights into the broader effects of myopia beyond its initial stages and evaluate the potential benefits of interventions that target the long-term management of myopia and its associated conditions, such as cataract, macular degeneration, open-angle glaucoma, and retinal detachment. Additionally, this survey will gather information on healthcare access, treatment costs, and reimbursement to provide a more comprehensive evaluation of the myopia economic burden.

## 2. Study objectives

This study aims to assess the impact of myopia in childhood and its long-term complications in adulthood on HRQoL, providing valuable data for cost-effectiveness models.

It will gather country-specific information on healthcare resource utilization, including treatment costs, access to care, and reimbursement policies. In addition, the study will examine how myopia and its treatments affect daily activities and evaluate treatment satisfaction. Ultimately, the data collected will be used to publish data on the impact of myopia on children’s quality of life, as well as inform health economic models. No data on the efficacy of myopia treatments will be collected in this study, nor will the occurrence of treatment-related adverse events be collected.

## 3. Study design

This is a digital, observational, cross-sectional survey conducted in the United Kingdom, France, Germany, Italy, Spain, Denmark, and Sweden. Additional comparable EU countries may be considered for inclusion in a second wave to ensure the recruitment rate is achieved.

Data will be collected from parents of children who are already diagnosed with myopia or from adults suffering from myopia, plus a long-term complication that is linked to increased risk of occurrence, namely cataract, macular degeneration, open-angle glaucoma, retinal detachment, or severe visual impairment/legal blindness.

The survey will include mainly closed-ended questions, as well as validated instruments. It is designed to minimize participant burden while maximizing the relevance and quality of the data collected. All participants will complete the survey once, online, on their computer or their mobile devices, and no longitudinal follow-up is planned.

## 4. Study population & sample size

Data will be collected among a minimum of 350 patients from hospitals across 7 countries: the United Kingdom, Germany, France, Italy, Spain, Denmark, and Sweden (Table 1). In each of the countries, a minimum of 50 participants will be recruited from an eligible study population using convenience sampling. Patients who meet the inclusion criteria and are willing to participate will be invited to take part in the study. Although the sampling will not be random, this approach is intended to ensure feasibility while targeting participants who meet predefined eligibility criteria.

**Table 1:** Distribution of minimum sample size across seven countries.

| Country | Minimum Sample Size |
| --- | --- |
| United Kingdom | 50 |
| Germany | 50 |
| France | 50 |
| Italy | 50 |
| Spain | 50 |
| Denmark | 50 |
| Sweden | 50 |
| Total patients | **350** |

The eligible study population consists of two groups:

- Parents (or other proxy) of children or adolescents aged 5 years or older suffering from myopia (low, moderate, or high myopia) defined by their refractive error.
  - Low myopia: from -0.5 D (inclusive) to -3.0 D (inclusive)
  - Moderate myopia: from -3.0 D to -6.0 D (inclusive)
  - High myopia: <-6.0 D.
- Adults suffering from myopia and one of the following myopia-related long-term complications. Patients where the complication is likely unrelated to myopia (e.g., cataract caused by diabetes) will be excluded.
  - Cataract
  - Macular degeneration
  - Open-angle glaucoma
  - Retinal detachment
  - Legal blindness/severe visual impairment.

To ensure the proper distribution of patients across the different groups, the total sample of at least 350 patients will be stratified based on the severity of myopia in children (low, moderate, high) as well as based on the presence of specific long-term complications. Table 2 provides an overview of the stratification of patients by myopia severity in childhood and associated long-term eye complications in adulthood. These amounts represent the optimal minimum number of patients for each category. Variation is expected per country.

**Table 2:**
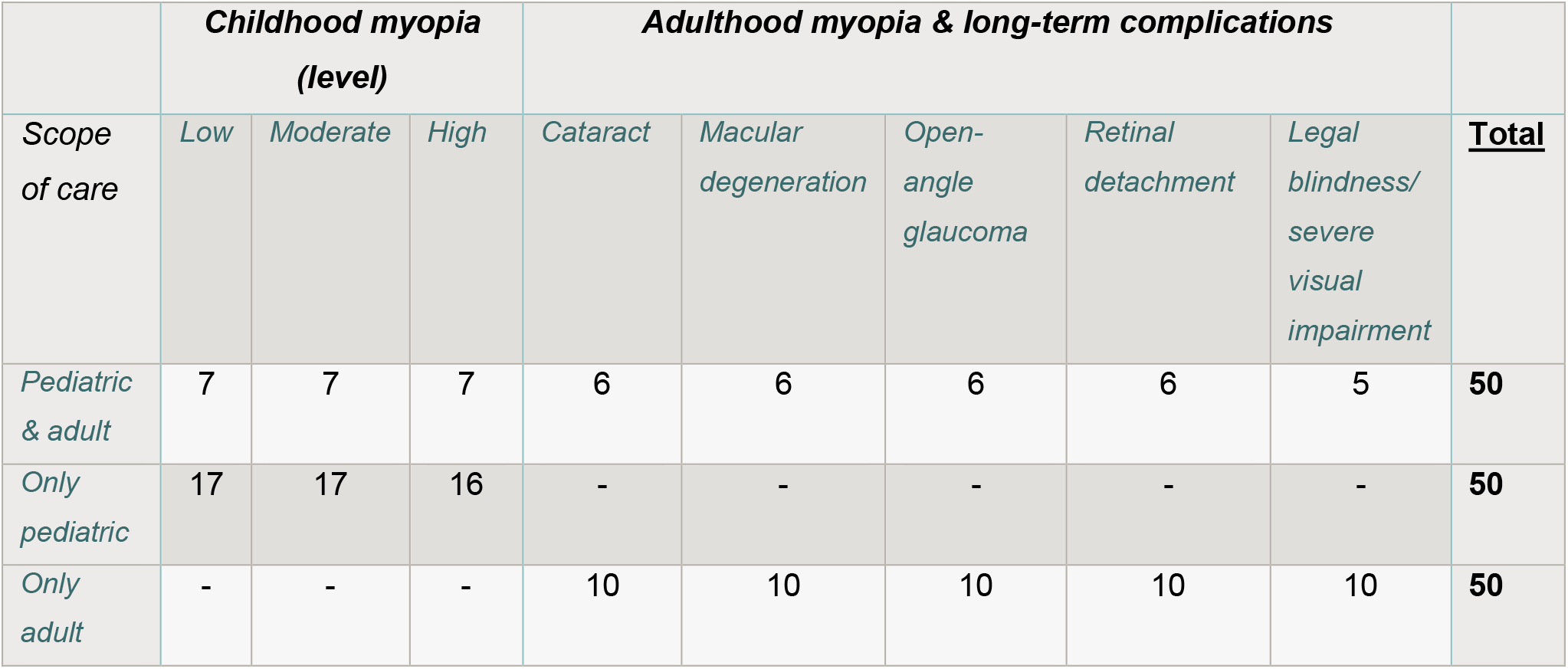
Stratification of patients by myopia severity in childhood and associated long-term complications in adulthood, across the different patient groups treated by the KOL (Key Opinion Leaders).

A minimum of 350 patients across 7 countries is the target recruitment goal. If there are multiple eye specialists (KOLs) per country, this number will increase by 50 per additional research sites. The stratification goals will be communicated clearly to the KOLs to ensure accurate enrolment of participants into the appropriate categories. To account for possible differences in recruitment between countries, flexibility is built into the recruitment process. If a country exceeds its recruitment target, the recruitment period may be extended to allow further participation and maximize data collection. In cases where a country does not reach the minimum number of participants, additional strategies will be implemented, such as extending the recruitment period or adapting local recruitment methods. The timeline may be adjusted accordingly to ensure that overall stratification goals are met.

## 5. Data collection

The survey will start with an online Informed Consent Form. Participants will be asked to indicate in which hospital they are being treated, as well as whether they are completing the questionnaire on behalf of a child or for themselves. Participants will be divided into two groups based on who is completing the questionnaire.

Group 1 respondents are parents (or other proxies) of children and adolescents suffering from myopia. Respondents will first fill out demographic questions, followed by a generic measure of HRQoL **EQ-5D-Y-3L** (proxy) with Bolt-on vision. Afterwards, participants will fill out myopia- or eye-specific questions on current myopia state and treatment, diagnosis and treatment, treatment side effects and satisfaction, healthcare resource utilization, and activity. Finally, participants will fill out the **PedEyeQ** (proxy), a measure of vision-related quality of life for children with myopia.

Group 2 respondents are adult patients suffering from myopia and a long-term complication of myopia. Respondents will first fill out demographic questions, followed by a generic measure of HRQoL **EQ-5D-5L** (self-complete version) with Bolt-on vision. Afterwards, participants will fill out myopia-or eye-specific questions on current myopia state, diagnosis and treatment, long-term complications, treatment and satisfaction, and healthcare resource utilization. Finally, participants will fill out the **NEI-VFQ-25**, a measure of vision-related quality of life for adults with myopia and a long-term complication.

The survey is expected to take approximately 12-15 minutes to complete by the proxy and 8-10 minutes by the adult participant. The participant only needs to complete the survey once. The full survey will be programmed in Qualtrics (version April 2025). All questions are optimized to be displayed on both desktop and mobile devices. To avoid having large quantities of missing data, respondents are required to answer all questions. Because of this, there will be no missing data, unless participants decide not to finish the survey.

### 5.1. Questionnaire

#### 5.1.1. Demographics

- **Age of the child** in years (between 5 and 17)
- **Sex of the child** (i.e., girl or boy)
- **Relationship of the person filling out the questionnaire on behalf of the child** (i.e, parent, grandparent, caregiver/guardian, relative)
- **Age of the proxy** in years
- **Sex of the proxy** (i.e., Male, female, prefer not to answer)
- **Age of the adult** in years
- **Sex of the adult** (i.e., Male, female)

#### 5.1.2. Myopia-and-eye-condition-specific questions

##### Group 1 – Proxy

- Initial diagnosis of myopia and the first treatment received by the child
- Child’s most recent eye measurements, the current treatment the child is receiving for myopia, and the effectiveness of these treatments.

##### Group 2 – Adults

- Initial diagnosis of myopia, including the degree of near-sightedness and treatment received at that time.
- Current myopia state, any eye-related complications experienced, the undergone treatment, and the feeling about the current eye condition.

#### 5.1.3. Healthcare Resources Utilization

Participants will answer questions on healthcare resource utilization (e.g., visits to the ophthalmologist), including treatment costs, whether treatment is reimbursed, and access to eye-care services.

#### 5.1.4. Activity questions

Consists of questions on the number of hours the child spends doing outdoor activities and up-close activities.

#### 5.1.5. Validated instrument: Vision-related Quality of Life and Functioning

- **PedEyeQ for children with myopia:** The PedEyeQ (Pediatric Eye Questionnaire) proxy version is an HRQoL assessment tool specifically designed to measure the impact of pediatric eye conditions on children. The tool consists of age-appropriate questionnaires (5-11 years old and 12-17 years old) that assess how vision problems affect daily activities, social interactions, school performance, and overall well-being of the child.
- **NEI-VFQ-25:** The NEI-VFQ-25 (National Eye Institute Visual Function Questionnaire-25) is a widely used tool to assess VRQoL in individuals aged 18 and older. It consists of 25 items that measure the impact of vision impairment on daily activities and well-being across 12 domains, including visual function, mental health, social functioning, and role limitations. The NEI-VFQ-25 helps evaluate how visual problems affect a person’s ability to perform tasks like reading, driving, and interacting socially.

#### 5.1.6. Validated instrument: Health-related Quality of Life and Functioning

- **EQ-5D-Y-3L (proxy):** The EQ-5D-Y-3L proxy version is an instrument used to assess the HRQoL of children and adolescents from the perspective of a parent, caregiver, or another proxy. It evaluates five dimensions of health—mobility, looking after oneself, doing usual activities, pain/discomfort, and feeling worried/sad—using a three-level scale (no problems, some problems, or a lot of problems), along with a visual analogue scale (VAS) from 0 (worst imaginable health) to 100 (best imaginable health)^9^. This version allows caregivers to report on the child’s health and estimate utility values. The EQ-5D-Y-3L country specific value sets (when available) will be used to calculate utility values.
- **Vision Bolt-Ons**: The vision bolt-on from EuroQol is an extension to the EQ-5D tool to specifically address the impact of vision impairment on a person’s HRQoL and consists of a single question in the same format as the EQ-5D-Y-3L instrument, with 3 levels.
- **EQ-5D-5L (self-complete version):** The EQ-5D-5L, a widely used instrument to value health in adults, includes five questions covering mobility, self-care, usual activities, pain/discomfort, and anxiety/depression. Each domain is rated from 1 (best quality) to 5 (worst quality), along with a VAS from 0 (worst imaginable health) to 100 (best imaginable health)^9^. The EQ-5D-5L country-specific value sets will be used to calculate utility values for adults with myopia and its complications.
- **Vision Bolt-on:** The vision bolt-on from EuroQol is an extension to the EQ-5D tool to specifically address the impact of vision impairment on a person’s quality of life and consists of a single question in the same format as the EQ-5D-5L instrument, with 5 levels.

## 6. Survey flow

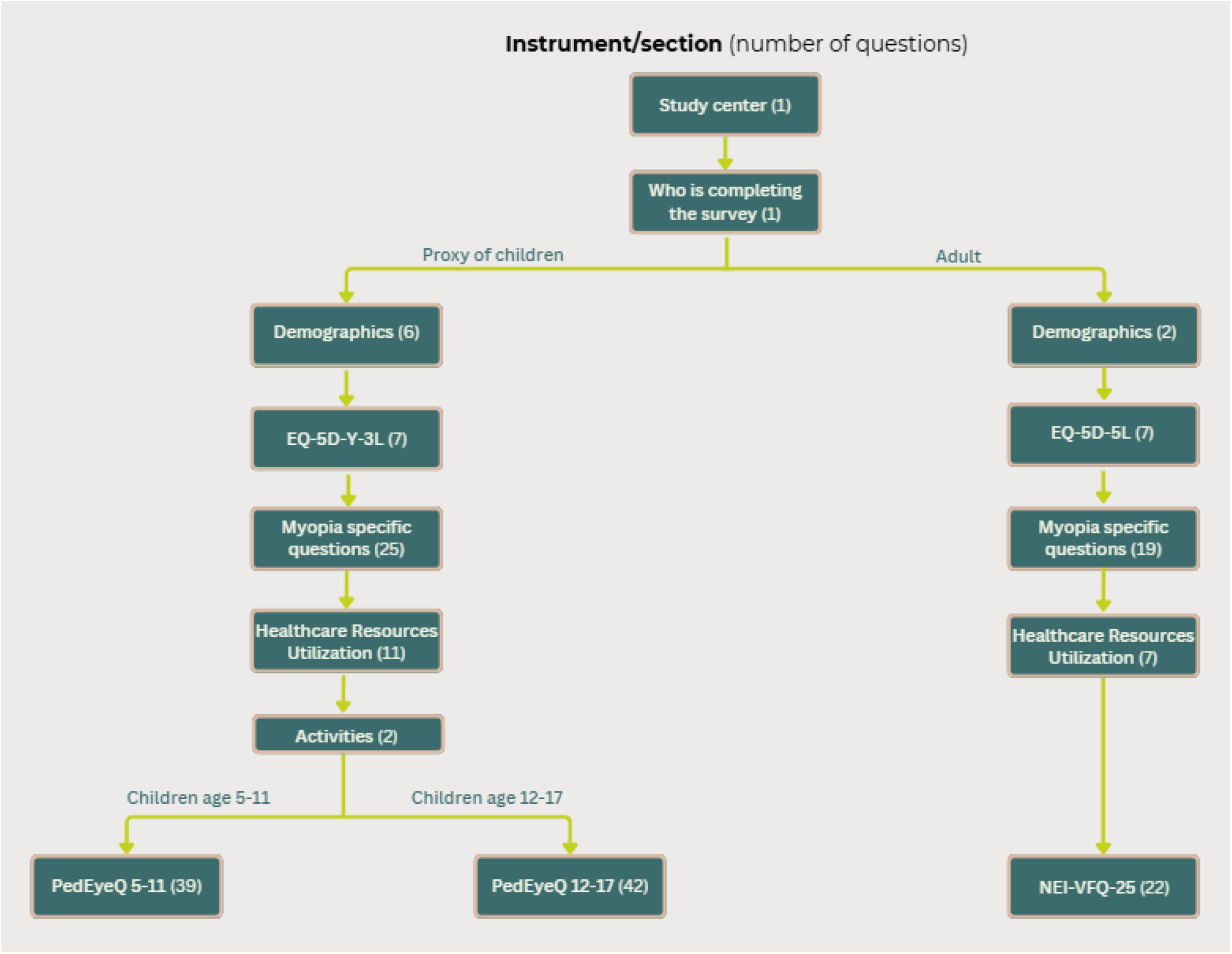

## 7. Analysis

The study is purely descriptive. No hypotheses are aimed to be tested. Descriptive statistical methods will be used to present the quality of life and medical resource utilization per myopia severity. Analysis techniques include the calculation of utilities, frequency tables for categorical variables, and means and distributions for continuous variables, univariable and multivariable regression, and resource utilization. It is planned to use the statistical analysis software R, SAS/STAT to analyse the collected data. Analyses include producing the following data tables:

- Distribution of patients, by myopia severity (low, moderate, high myopia), per country, and overall
- Distribution of age and gender, by myopia severity (low, moderate, high myopia)
- Distribution of utility values, by myopia severity (low, moderate, high myopia)
- Distribution of myopia progression rate, by myopia severity (low, moderate, high myopia)
- Distribution of treatment types received at diagnosis and current treatments, per country and overall
- Distribution of side effects experienced from current treatment(s)
- Number of hours a child spends doing outdoor activities and doing near-work tasks, by myopia severity (low, moderate, high myopia)
- Acceptability with current disease status
- Healthcare resource utilization: difficulties accessing eye care services, reimbursement of treatment, frequency of eye doctor visits, out-of-pocket costs on vision care, per country, and overall

## 8. Survey conduct & procedures

### 8.1. Recruitment

Key Opinion Leaders (KOLs) from participating hospitals will invite eligible patients to participate in the survey during their scheduled follow-up visits. The KOLs will recruit participants by distributing a pamphlet containing a QR code. Scanning the QR code will direct participants to the survey. The online survey will begin with a digital informed consent form (ICF) to confirm the patient’s willingness to participate and obtain electronic consent.

The KOLs will not have access to the participants’ responses, and the survey responses will not be traceable to the individual participants by the researchers. Participation in the survey will not affect the patient’s treatment in any way.

Participants will be informed that their participation is voluntary, and they may withdraw at any time without consequence. The study is purely observational without any medical intervention, drug, or medical device administration.

### 8.2. Incentive

Participating hospitals will receive a centre fee estimated at €1500 per centre, paid in a fixed and a variable part: €500 fixed honoraria per participating centre, plus €20 for the study centre per patient who fully completed the questionnaire.

Respondents can, in return for their participation in the survey, claim a €20 voucher with a variety of options to claim the rewards based on the country of the participant.

### 8.3. Monitoring of data quality

To ensure the integrity, accuracy, and completeness of the data, regular checks will be conducted throughout the data collection process. These checks will focus on identifying any discrepancies between responses from the same patient, will remove speeders, will highlight missing responses or early termination of the survey, or identify bots. Regular feedback on data collection progress will be provided to the KOLs, allowing any issues to be addressed in real-time, ensuring high data quality and reliability throughout the study.

## 9. Data storage, security, ownership, and protection

Data will be collected through the Qualtrics platform and securely stored on servers managed by CHEOPS, a dedicated IT-security company (CHEOPS.com). The data will be stored for 7 years on a cloud server managed by Services in Health Economics (SHE) BV, and continuously monitored by CHEOPS, in accordance with the General Data Protection Regulation (GDPR) and the Network and Information Security Directive 2 (NIS-2) (cybersecurity) regulations. Only authorized researchers will have access to raw data. SHE BV is the sole owner and processor of all collected data in this study.

## 10. Timelines

It is planned to start data collection in June 2025 and end it when all data have been collected, with an estimated duration of 3 to 6 months. Initial results in table format are expected to be shared with the study sponsors and participating centres by Q4 of 2025.

## 11. Publication

Survey results will be published in peer-reviewed articles, aiming towards high-impact journals.

## Data Availability

This article describes a study protocol, no datasets are associated with this protocol.

## 13. Ethical considerations

Individuals will be able to complete the online survey only after agreeing to the online consent form. The survey is completely anonymous: no names or other personal identifiers (e.g., email addresses, initials, postal address, date of birth, national registration number, medical file number, IP address, location data, etc.) will be recorded or included in any database.

This study protocol and the informed consent documents will be submitted for review and approval by the appropriate IRB/IEC before the study may begin. The study protocol has been submitted and approved by the appropriate IECs in Germany, Sweden, Denmark, Italy, and Spain. Additionally, the study has been submitted on ClinicalTrial.gov under the following ID: NCT06912802. Any amendments to the study protocol (e.g., any expansion to additional countries) will be submitted for re-evaluation and approval by the IRB/IEC prior to implementation. The Principal Investigator has completed training in Good Clinical Practice (GCP), in accordance with the International Conference on Harmonisation (ICH) guidelines.

The study is purely observational, it will not entail any medical intervention, drug, or medical device administration. The study will be conducted in accordance with the principles contained in the Declaration of Helsinki (updated version), the Convention on Human Rights and Biomedicine, and the laws relating to transparency, the prevention of corruption, and the current data protection regulations.

## References

1. Cooper J, Tkatchenko A V. A Review of Current Concepts of the Etiology and Treatment of Myopia. Eye Contact Lens. 2018;44(4):231–247. doi:10.1097/ICL.0000000000000499

2. Németh J, Tapasztó B, Aclimandos WA, et al. Update and guidance on management of myopia. European Society of Ophthalmology in cooperation with International Myopia Institute. Eur J Ophthalmol. 2021;31(3):853–883. doi:10.1177/1120672121998960

3. Bullimore MA, Ritchey ER, Shah S, Leveziel N, Bourne RRA, Flitcroft DI. The Risks and Benefits of Myopia Control. Ophthalmology. 2021;128(11):1561–1579. doi:10.1016/J.OPHTHA.2021.04.032

4. Wu PC, Chuang MN, Choi J, et al. Update in myopia and treatment strategy of atropine use in myopia control. Eye (Lond). 2019;33(1):3–13. doi:10.1038/S41433-018-0139-7

5. Morgan IG, Ohno-Matsui K, Saw SM. Myopia. The Lancet. 2012;379(9827):1739–1748. doi:10.1016/S0140-6736(12)60272-4/ATTACHMENT/0F5F6EBF-EF8E-40A2-976A-151807B53A81/MMC1.PDF

6. Azuara-Blanco A, Logan N, Strang N, et al. Low-dose (0.01%) atropine eye-drops to reduce progression of myopia in children: a multicentre placebo-controlled randomised trial in the UK (CHAMP-UK)-study protocol. Br J Ophthalmol. 2020;104(7):950–955. doi:10.1136/BJOPHTHALMOL-2019-314819

7. Salomon JA, Haagsma JA, Davis A, et al. Disability weights for the Global Burden of Disease 2013 study. Lancet Glob Health. 2015;3(11):e712–e723. doi:10.1016/S2214-109X(15)00069-8

8. Cui Z, Zhou W, Chang Q, et al. Cost-Effectiveness of Conbercept vs. Ranibizumab for Age-Related Macular Degeneration, Diabetic Macular Edema, and Pathological Myopia: Population-Based Cohort Study and Markov Model. Front Med (Lausanne). 2021;8. doi:10.3389/FMED.2021.750132

9. Herdman M, Gudex C, Lloyd A, et al. Development and preliminary testing of the new five-level version of EQ-5D (EQ-5D-5L). Qual Life Res. 2011;20(10):1727–1736. doi:10.1007/S11136-011-9903-X

